# Rapid emergence and transmission of virulence-associated mutations in the oral poliovirus vaccine following vaccination campaigns

**DOI:** 10.1101/2023.03.16.23287381

**Authors:** Katharine S. Walter, Jonathan Altamirano, ChunHong Huang, Yuan J. Carrington, Frank Zhou, Jason R. Andrews, Yvonne Maldonado

## Abstract

There is an increasing burden of circulating vaccine-derived polioviruses (cVDPVs) due to the continued use of oral poliovirus vaccine (OPV). However, the informativeness of routine OPV VP1 sequencing for the early identification of viruses carrying virulence-associated reversion mutations has not been directly evaluated in a controlled setting. We prospectively collected 15,331 stool samples to track OPV shedding from vaccinated children and their contacts for ten weeks following an immunization campaign in Veracruz State, Mexico and sequenced VP1 genes from 358 samples. We found that OPV was genetically unstable and evolves at an approximately clocklike rate that varies across serotypes and by vaccination status. Alarmingly, 28% (13/47) of OPV-1, 12% (14/117) OPV-2, and 91% (157/173) OPV-3 of Sabin-like viruses had ≥1 known reversion mutation. Our results suggest that current definitions of cVDPVs may exclude circulating virulent viruses that pose a public health risk and underscore the need for intensive surveillance following OPV use.

## Introduction

The global use of oral poliovirus vaccine (OPV), a live, attenuated virus, has been essential in reducing the burden of poliomyelitis caused by wild poliovirus by 99.9% since 1988^1,2^. The vaccine covers all three wild poliovirus serotypes, is inexpensive, easy to deliver, and—because it is a live virus—is transmissible from vaccinated children onwards, extending the reach of vaccination campaigns. However, OPV vaccination is risky—the vaccine virus is genetically unstable and continues to evolve in vaccinated individuals and their contacts. If OPV mutates at positions responsible for attenuation of the vaccine virus, it can regain virulence. OPV can regain virulence within a vaccinated individual, causing vaccine-associated paralytic poliomyelitis, though rarely^3^. OPV may also revert to virulence over transmission chains, generating outbreaks of circulating vaccine derived polioviruses (cVDPVs).

To reduce the risks of cVDPVs, in May 2016, trivalent OPV vaccines (containing serotypes 1, 2, and 3) were replaced with a bivalent OPV (containing serotypes 1 and 3), following World Health Organization recommendations. The WHO recommended that routine immunization with at least one dose of inactivated poliovirus vaccine (IPV) prior to the switch to bivalent OPV, to increase protection against paralytic disease^4^. However, access to IPV was limited, leaving large populations of children immunologically naïve to poliovirus type 2. Since the global switch to bivalent OPV, the majority of paralytic poliovirus cases are now caused by cVDPVs, not wild poliovirus, the majority of which are serotype 2 viruses (cVDPV2). From 2016 to 2022, type 2 cVDPVs emerged 68 times in 34 countries, leading to more than 1500 cases of paralytic polio^5^ (Fig. 1).

**Figure 1.**
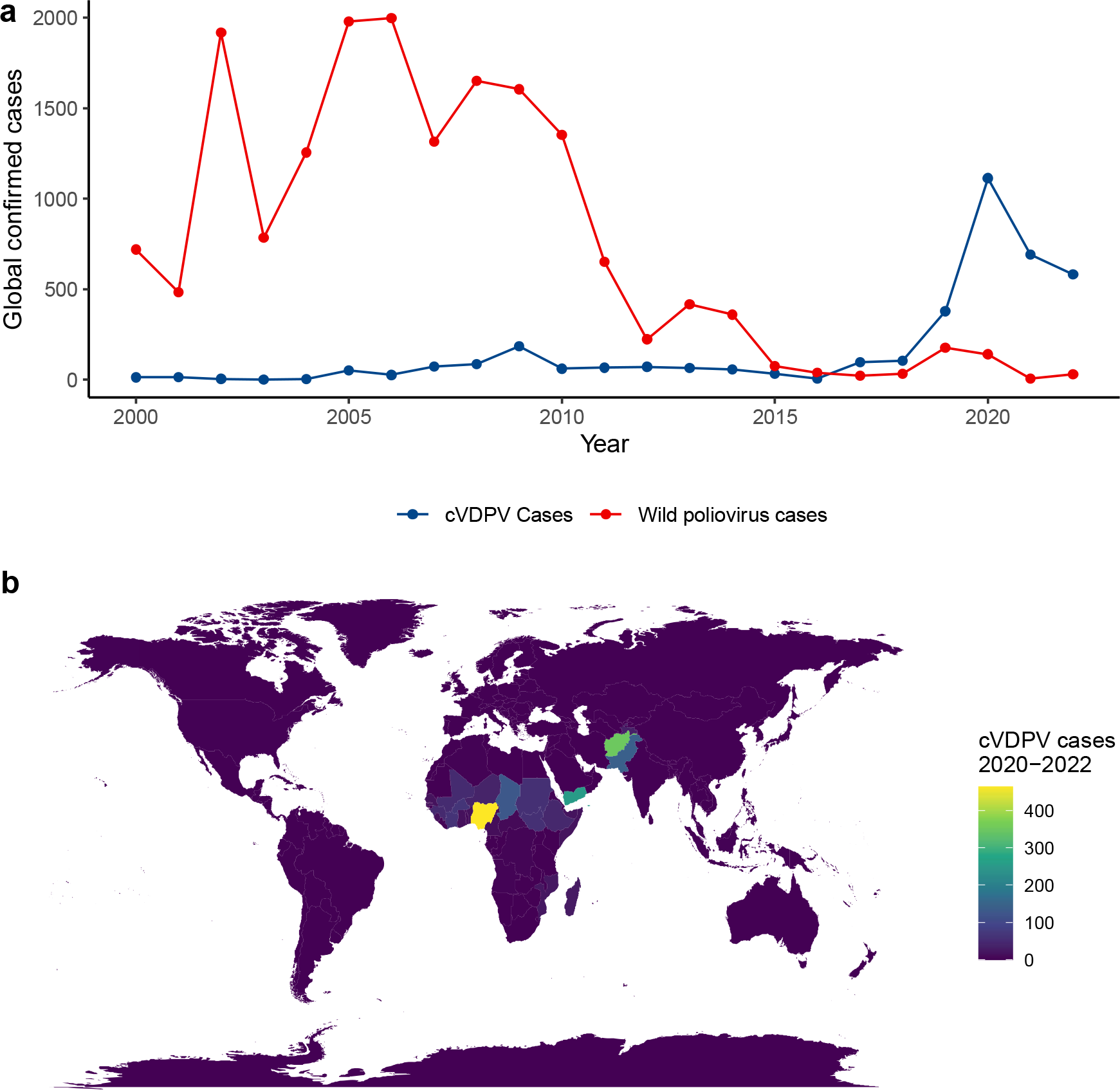
The increasing global burden of vaccine-derived poliovirus. (a) Confirmed cases of paralytic polio reported to the World Health Organization from 2000-2022. Color indicates source: circulating vaccine derived poliovirus (cVDPV) and wild poliovirus cases. (b) Map of countries reporting cVDPV cases in from 2020-2022; fill indicates number of cVDPV cases identified. Data from the World Health Organization.

The current public health response to outbreaks of cVDPV type 2 is vaccination with monovalent OPV-2 (mOPV-2), to increase population-level immunity and reduce transmission risk. However, the outbreak response itself is risky, as mOPV-2 use can generate new cVDPV outbreaks.

Outbreak responses are guided by routine testing of cases of acute flaccid paralysis in children less 15 and environmental surveillance in more than 30 high-risk countries and sequencing of the poliovirus VP1 gene^6^. The Global Polio Eradication Initiative (GPEI) defines VDPVs by their divergence from OPV in the VP1 gene^6^. cVDPVs are defined by evidence of VDPV transmission, through genetically linked viruses from individuals with acute flaccid paralysis or environmental samples^6^ and assigned to new or existing genetic clusters^7,8^. The identification of new cVPDVs results in an epidemiological investigation and an immunization response^6^.

Previous genomic studies of type 2 cVDPVs and OPV-2 shedding in vaccinated children and contacts have identified strong within-host selection and tight transmission bottlenecks that govern the early evolution of type 2 viruses away from the Sabin vaccine strain along a predictable pathway to virulence^11,12^. Yet previous studies have not yet integrated detailed molecular and epidemiological data to assess the use of VP1 sequencing for the early identification of viruses posing a public health risk. Given that global surveillance and outbreak responses rely on VP1 sequencing, we need a better understanding of how informative VP1 diversity is for characterizing risk of reversion to virulence and transmission of virulent viruses.

Here, we followed the evolution and transmission of the three OPV serotypes during a prospective study of OPV shedding following a vaccine campaign in semi-rural communities in Veracruz State, Mexico^13^. We sequenced OPV VP1 genes from stool sampled from vaccinated children, their household contacts, and community members in unvaccinated households for 10 weeks following the vaccination campaign. This provides a rare opportunity to observe the early evolution and transmission of the three OPV serotypes following vaccination campaigns and to evaluate the current use of VP1 sequencing as an epidemiological tool.

## Methods

### Prospective sampling

We conducted a prospective study of OPV shedding and transmission following a trivalent OPV vaccination campaign during Mexico’s February 2015 National Health Week in three semi-rural populations in Orizaba, Veracruz State^13^ (Fig. 2). Wild poliovirus has not been reported since 1994 in Mexico. Mexico’s national vaccine program has included routine IPV as well as two annual OPV vaccination campaigns since 2007 and therefore provides a model for transmission of OPV in a setting with routine IPV immunization, an epidemiological context which will increasingly exist as IPV replaces OPV and OPV is only used as a tool for cVDPV or wild poliovirus outbreak response.

**Figure 2.**
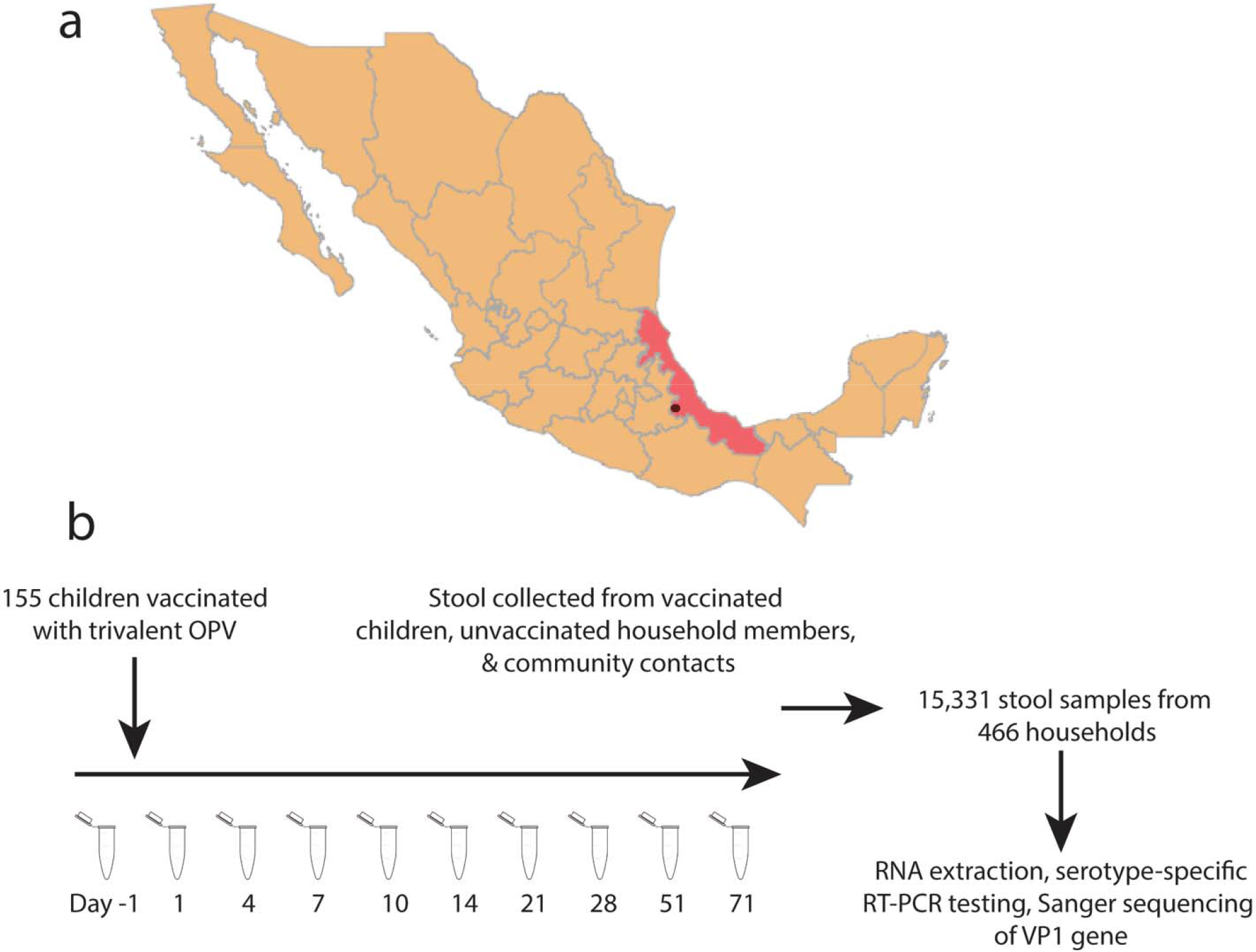
Prospective study of OPV shedding among vaccinated children, household members, and community members following a vaccination campaign. (a) Map of Mexico with Veracruz State highlighted and Orizaba city, where the study took place, indicated with a red point. (b) We conducted a prospective observational study of OPV viral shedding following a vaccination campaign. 155 children were enrolled and vaccinated with trivalent OPV. Stool was collected from vaccinated children, unvaccinated household members, and community members in unvaccinated households. We extracted RNA from stool samples, tested samples with OPV serotype-specific RT-PCR, and Sanger sequenced positive samples.

We randomized 466 households to the vaccine and no-vaccine arm of our study across three communities which were assigned to OPV vaccination rates of 10%, 30%, and 70%. In total, 155 children (8.5% of the population) were vaccinated with trivalent OPV during the National Health Week. Vaccinated children and their household contacts were followed prospectively and stool was collected from all members of sampled households 1 day prior to the National Health Week OPV immunization campaign, and 1, 4, 7, 10, 14, 21, 28, 51, and 71 days following, for a total of 10 samples per person^13^. Study nurses collected immunization histories and additional demographic and clinical metadata from all study participants.

The study was approved by the Stanford University Institutional Review Board (Protocol #31546), the Comité de Etica, Bioseguridad e Investigación of the Instituto Nacional de Salud Pública (CI: 1260, No. 1581), and the Instituto Veracruzano para la Formación e Investigación en Salud (SESVER/IVEFIS//SIS/DIB/0109/02014, classification 15S).

### Sanger sequencing of VP1 gene

We extracted RNA from frozen stool samples and tested samples for OPV with serotype-specific RT-qPCR^13,14^. We Sanger sequenced the VP1 gene using the Y7 and Q8R primers^15^ and generated consensus sequences by aligning forward and reverse sequences to serotype-specific reference genomes in *Geneious* v7.

### Bioinformatic analysis

We aligned VP1 gene sequences of each OPV serotype to the corresponding Sabin vaccine sequence (GenBank Accession Numbers: AY184219, AY184220, AY184221) with MAFFT v.7^16^, with the algorithm recommended for closely related viral sequences (https://mafft.cbrc.jp/alignment/software/closelyrelatedviralgenomes.html), preserving the alignment length of the reference sequence. We used the default alignment strategy and a nucleotide sequence scoring matrix of 1PAM / κ=2, recommended for closely related DNA sequences.

We quantified pairwise distances between samples with the R package *ape* (pairwise deletion = TRUE)^17^. For household contacts and community members who were not vaccinated with OPV, we measured time since vaccination as the time since vaccination of the household member or the earliest vaccination given in the community, respectively. We visualized haplotype networks built from minimum spanning trees with the R package *pegas*^18^.

### Data availability

The full-length VP1 sequences used in our study are publicly available on GenBank (Submission 2683280, 2683353, and 2683683). The references sequences used for sequence alignment are publicly available on GenBank AY184219, AY184220, AY184221). Multiple sequence alignments and metadata to reproduce figures and analyses are publicly available on GitHub (https://github.com/ksw9/opv-consensus).

### Code availability

Code for analysis and figures is publicly available on GitHub (https://github.com/ksw9/opv-consensus).

### Global poliovirus data

We accessed World Health Organization poliovirus case data from the WHO extranet (https://extranet.who.int/polis/public/CaseCount.aspx), including both global and country-level reported cases from 2000-2022.

## Results

### Sample collection

During a prospective study of OPV shedding following a National Health Week vaccination campaign in Veracruz State, Mexico, we collected 15,331 stool samples over 10 weeks following vaccination. From these samples, 551 samples were positive for at least one OPV serotype, including 267 OPV-1, 402 OPV-2, and 317 OPV-3 positive samples. We sequenced 358 high quality VP1 genes from 174 individuals, including 18.4% (49/267) positive OPV-1 samples, 33.8% (136/402) positive OPV-2 samples, and 54.5% (173/317) OPV-3 samples. Sequence data was available for 99 vaccinated children, 41 household contacts of vaccinated children, 31 members of the community outside of vaccinated households, and 3 individuals without known vaccination status. For 84 infections (unique individuals and serotypes), longitudinal samples were available at 2-5 sampling points. The prospective study was conducted in three semi-rural villages; samples include 106 individuals from Capoluca, where 70% of children less than five in participating households were vaccinated; 49 from Campo Grande, where 30% were vaccinated; and 19 individuals from Tuxpanguillo, where 10% were vaccinated.

### Genetic instability of OPV

We observed a rapid rise of VP1 diversity in all three serotypes over weeks following OPV vaccination. The most common haplotype for both OPV-1 and OPV-2 in each sampling location represents the Sabin vaccine strain (Fig. 3), with a star-like pattern in the haplotype networks, where most haplotypes are directly connected to the Sabin haplotype. Outside of the dominant haplotype, most haplotypes are represented by a single sample.

**Figure 3.**
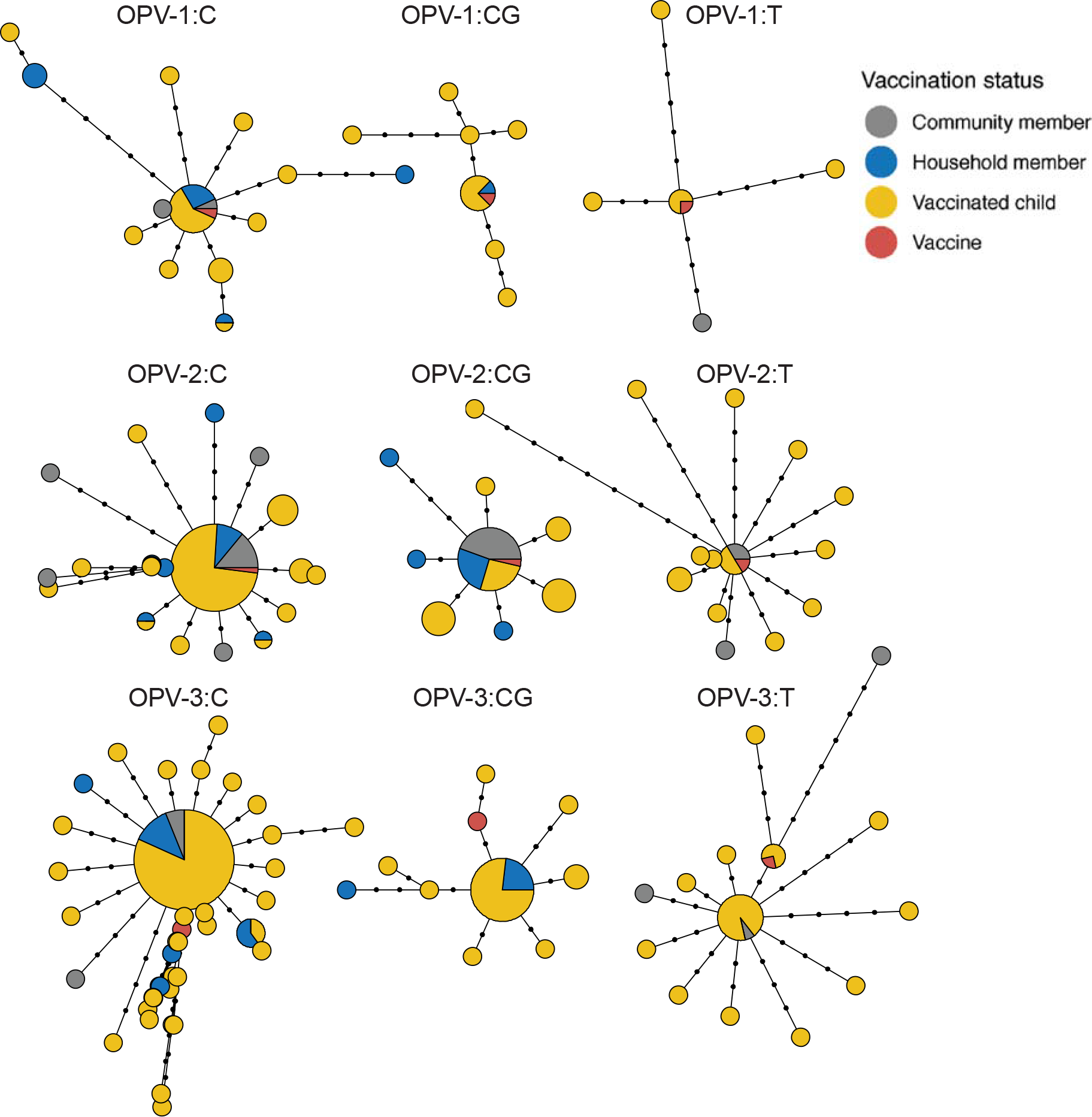
Limited overall genetic diversity in sampled OPV viral capsid protein 1 (VP1) gene following vaccination campaigns. For each OPV serotype and sampling site, haplotype networks of OPV VP1 representing sampled genetic diversity. Nodes indicate identical haplotypes (VP1 consensus sequences) and node size indicates the number of samples sharing a haplotype. Points on edges between nodes indicate the SNP distance between haplotypes and node colors indicate individual’s vaccination status. Labels indicate OPV serotype and study site. Site C, Capoluca, had 70% vaccination coverage of eligible children; site CG, Campo Grande: 30%; site T, Tuxpanguillo: 10%. One sample with a distant haplotype (>14 SNPs distant to the Sabin vaccine) was removed from OPV-2 site C, to aid visualization.

In contrast, for OPV-3, the dominant haplotype differs by a single nucleotide polymorphism (SNP) from the vaccine strain, a previously described attenuating site, C2493U (Fig. 3). Again, there is a star-like pattern of OPV-3 diversity, with many branches stemming directly from the dominant haplotype. As with OPV-2 and OPV-3, most haplotypes outside the dominant haplotype are represented by a single sample.

### Clock-like early evolution of OPV following vaccination campaigns

The Global Polio Eradication Initiative (GPEI) defines VDPVs by their divergence from the Sabin vaccine in the VP1 gene (>1% divergent or ≥10 substitutions for serotypes 1 and 3, 0.6% divergent of ≥6 substitutions for serotype 2)^6^. The majority, 99% (337/340), of all samples sequenced following vaccination fell under the thresholds for a VDPV. However, two samples from vaccinated children, collected 4 days after vaccination met the definition of a VDPV2 (VP1 genetic distance 14 and 8 SNPs from Sabin 2) and one sample from a household contact sampled 30 days following vaccination met the definition of a VDPV1 (10 SNPs distant from Sabin 1) (Fig. 4).

**Figure 4.**
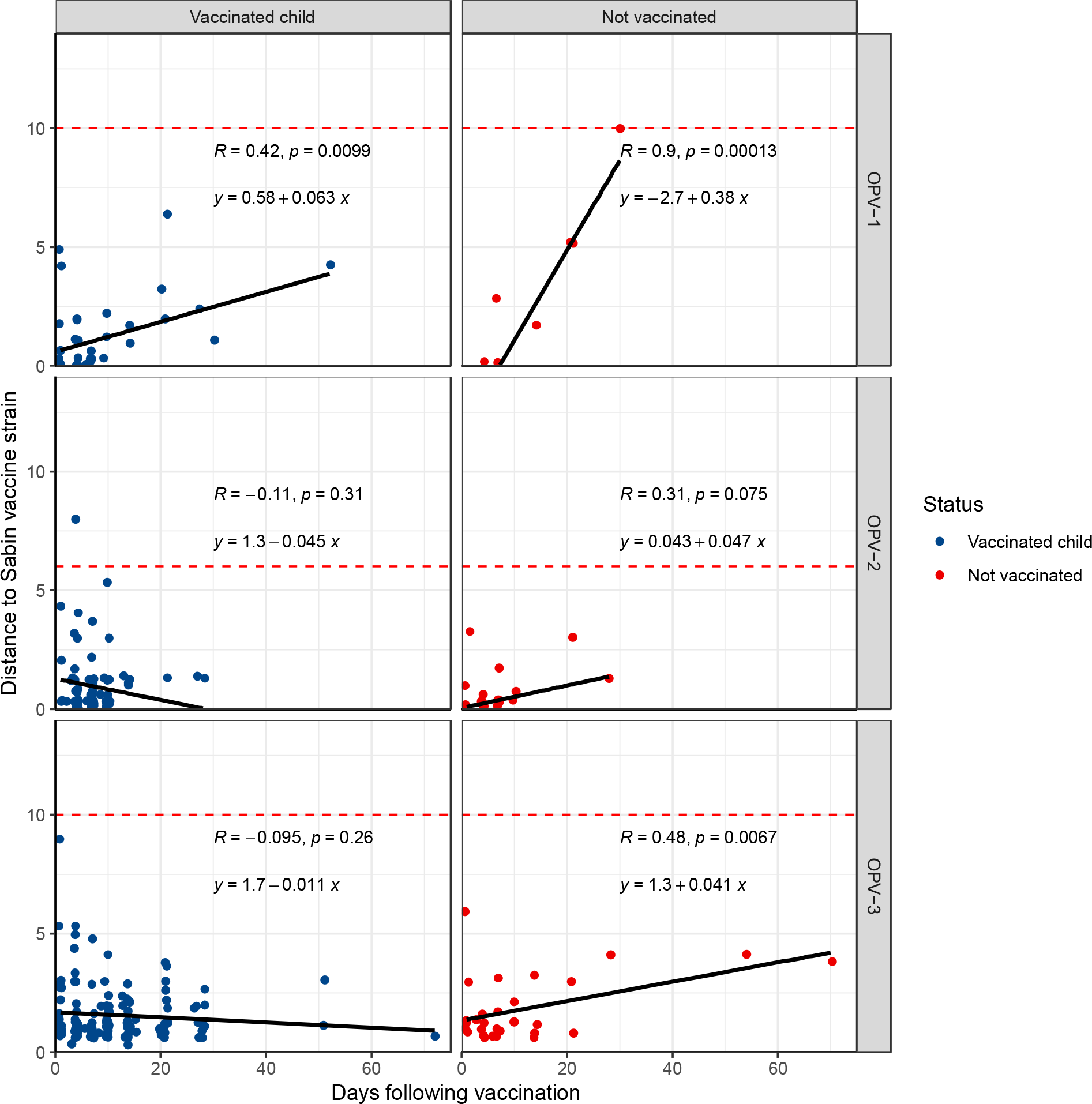
Measurable evolution in the OPV VP1 gene following vaccination campaigns. Genetic distance of the OPV VP1 gene to the Sabin vaccine strain versus time from vaccination for vaccinated children (blue) and not vaccinated study participants, including household contacts and unvaccinated community members (red), for each OPV serotype (a-c). Black lines and grey shading of a linear model for distance to the Sabin strain. Days following vaccination is measured as days from first vaccination (for vaccinated children), day from first household vaccination (for non-vaccinated household members), and days from first community vaccination (for non-vaccinated community members). Red dashed lines indicate the genetic distance threshold for a cVDPV: 10 SNPs distant from the Sabin vaccine for serotypes 1 and 3 and 6 SNPs distant for serotype 2.

We then examined if the early evolution of OPV follows a molecular clock and, therefore, if OPV divergence from Sabin vaccine strains could be used to estimate the duration of viral circulation. Genetic divergence of OPV-1 was significantly associated with time following vaccination in both vaccinated children (p < 0.001, Pearson’s r = 0.42) and non-vaccinated individuals(p < 0.001, Pearson’s r = 0.90) (Fig. 4). When considering all samples together, the observed mutations in OPV-1 corresponded to a VP1 gene evolutionary rate of 1.2 × 10^−4^ substitutions per site per day. Genetic divergence of OPV-2 was not significantly associated with time following vaccination in vaccinated children (p=0.3, Pearson’s r = -0.11) and was positively correlated with time among non-vaccinated individuals, though not significantly so (p =0.07, Pearson’s r = 0.31) (Fig. 4). OPV-3 divergence was positively associated with time following vaccination for non-vaccinated individuals (p = 0.007, Pearson’s r = 0.48), but not vaccinated children (p = 0.26, Pearson’s r = -0.1)

We reasoned that mutations at known attenuating positions may be under strong selection and could affect an estimate of molecular clock. However, excluding the known attenuating sites in the VP1 gene did not substantially change observed associations between genetic divergence and time (Fig. S1).

### Moderate signal of recent transmission in OPV VP1 gene sequence diversity

VP1 genetic diversity is also used by the GPEI to determine if a VDPV is genetically linked to previously collected environmental or clinical samples, which makes it a cVDPV, and whether viruses are linked to a previously identified cVDPV emergence or constitute a new emergence.

To test whether VP1 sequence divergence is genetically structured and could provide evidence of recent transmission, as is done with several pathogens, we compared pairwise genetic distances across samples. As expected with the relatively short duration of sampling following vaccination campaigns, 28.9% (694/2400) of OPV-1; 40.6% (67,16/16,504) of OPV-2; and 42.0% (12,498/29,974) of OPV-3 VP1 pairs of sequences were identical. Mean pairwise distance was 2.4 SNPs for OPV-1, 1.5 SNPs for OPV-2, and 1.4 SNPs for OPV-3 (Fig. 5).

**Figure 5.**
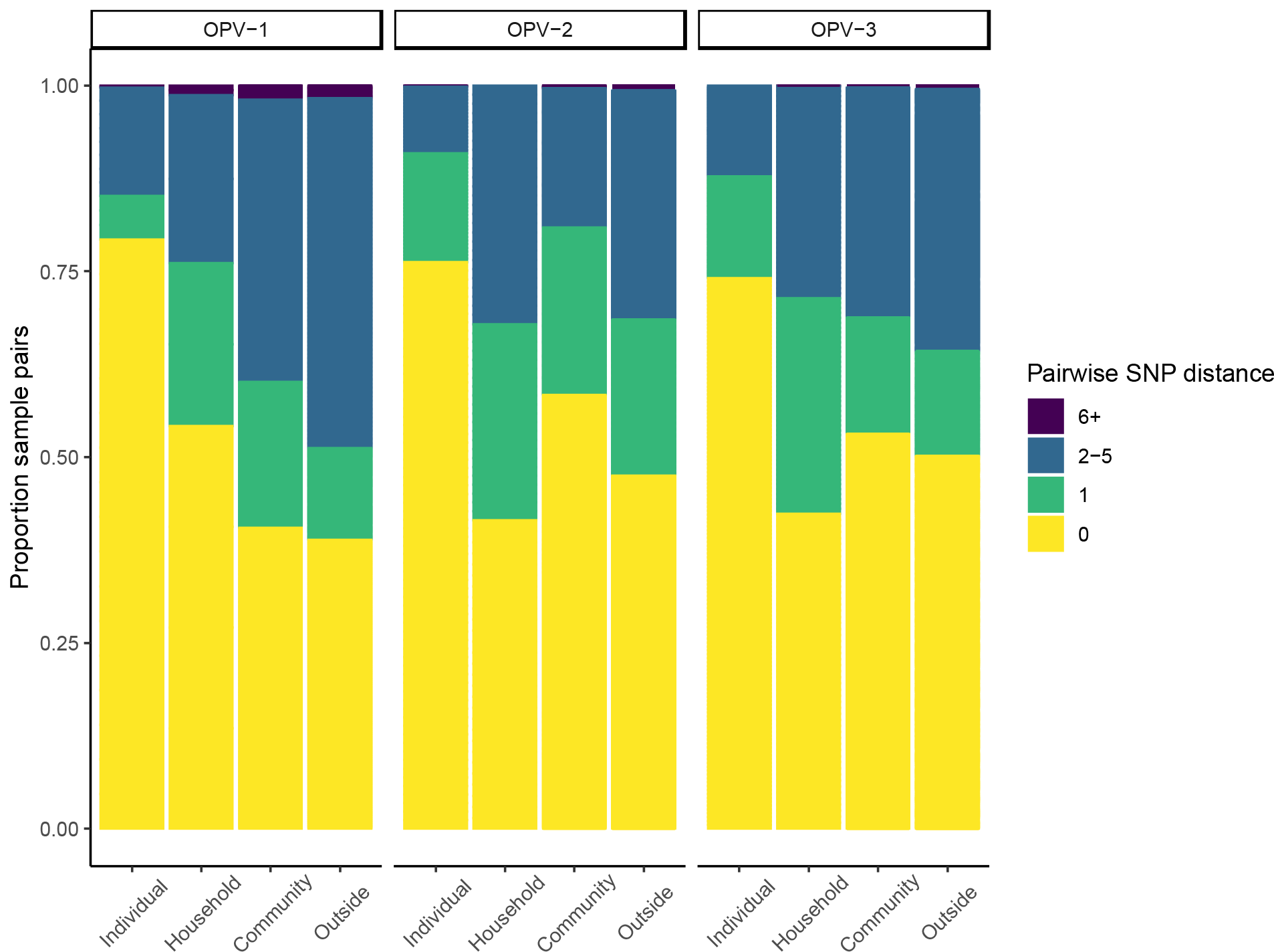
Moderate genetic structure of OPV VP1 gene from samples shed two months following vaccination campaigns. The proportion of pairs of OPV VP1 sequences within a given, binned pairwise genetic distance for samples collected longitudinally from the same individual, from the same household, the same community, or outside the community. Facets indicate OPV serotype.

In a general linear model, pairwise genetic distance between VP1 samples was elevated for individuals from the same household (aOR: 1.29; 95% CI: 1.11-1.50), from the same community (aOR: 1.49; 95% CI: 1.34-1.64), and from different communities (aOR: 1.63; 95% CI: 1.48-1.80), compared to samples collected from an individual. As expected, longitudinal samples from an individual were mostly conserved over the sampling time. Genetic structure varied across serotypes: compared to OPV-1, we observed decreased pairwise genetic distances for both OPV-2 (aOR: 0.64; 95% CI: 0.62-0.65) and OPV-3 (aOR: 0.58; 95% CI: 0.57-0.60).

### Rapid loss of OPV attenuating mutations in the VP1 gene following vaccination campaigns

We then examined known mutations at key attenuating sites in the vaccine virus, associated with the reversion of vaccine viruses to virulence. Of three known virulence attenuating substitutions in the OPV-1 VP1 gene, 16% (8/49) of samples had a A2749G (Sabin to virulent wildtype) mutation, including 4 vaccinated children and 2 household contacts. The mutation first appeared 14 days following vaccination in a vaccinated child and contact in one house and in another vaccinated child independently and rose in frequency, although there were limited OPV-1 samples available 4+ weeks after vaccination. It was observed in stool up to 52 days after the vaccination campaign. A single sample shed by a vaccinated child harbored a A2795G substitution, and no U2879C attenuating mutations occurred (Fig. 6).

**Figure 6.**
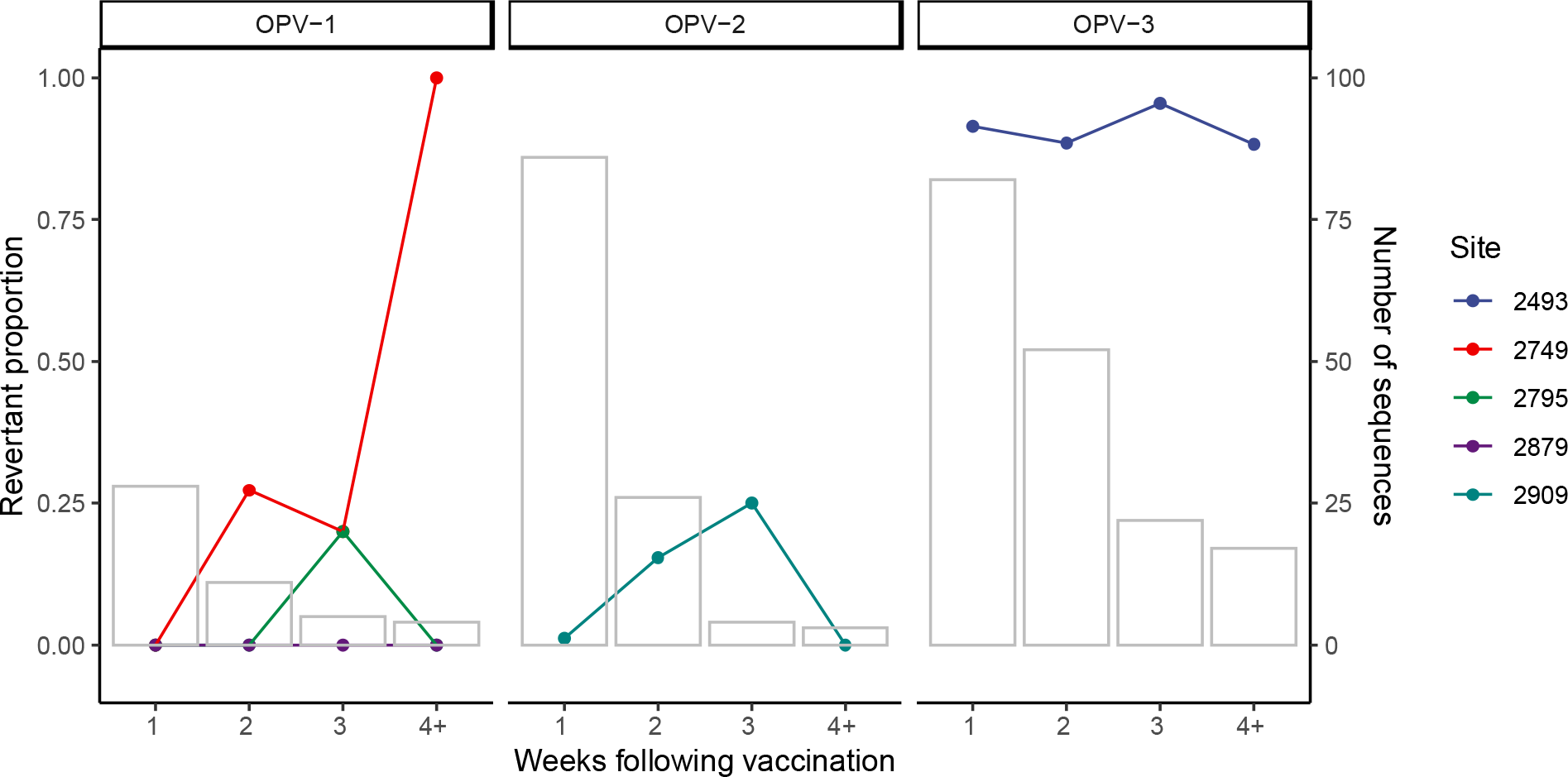
Loss of OPV attenuating mutations following vaccination. For each serotype, points indicate the revertant proportion, the proportion of samples with a mutation at a key attenuating position in OPV in the weeks following vaccination. Bars indicate the total number of sequences available for that sampling week. Each serotype has unique attenuating sites.

Among OPV-2 samples, the known VP1 reversion mutation, U2909C/A, occurred in 4.4% (6/136) of samples, in four vaccinated children. The substitution was first observed 7 to 14 days after vaccination; two children shed OPV-2 carrying this reversion mutation on multiple days. The mutation was observed in stool up to 21 days following the vaccination campaign, though again, there were limited OPV-2 samples available 4+ weeks after vaccination. No individuals had mutations present at the adjacent site 2908, also associated with reversion^19^.

Among OPV-3 samples, the majority, 91% (157/173), had a C2493A/G/U eversion substitution and an additional 11 samples had a mixed allele or no allele call at that site, possibly evidence of mixed infection. Samples with a C2493A/G/U mutation included 95% (89 of 94) of individuals with OPV-3 sequences available, including 63 vaccinated children, 18 household members, and 8 individuals outside of vaccinated households. The mutation first appeared one day following the vaccination in vaccinated children, household members, and individuals outside vaccinated households and it was observed in stool up to 72 days after the vaccination campaign from a vaccinated child and 70 and 54 days from virus shed in a household member and community member, respectively.

## Discussion

Global poliovirus surveillance uses molecular epidemiology to directly guide outbreak responses. However, the informativeness of the routinely sequenced marker gene, VP1, in the early identification of VDPVs posing a public health risk has not been evaluated in a controlled setting.

Here, we leveraged OPV sampled for 10 weeks following an immunization campaign in a population previously vaccinated with IPV to assess the epidemiological information within VP1 samples. We found that all three OPV serotypes are genetically unstable in the weeks following vaccination campaigns, consistent with previous reports. We identified evidence of clocklike VP1 evolution across serotypes (though this signal differed by vaccine status) and significant genetic structure. However, we found evidence of frequent reversion substitutions among viruses classified as Sabin-like, suggesting that current definitions of cVDPVs may exclude circulating viruses that have regained key virulence-associated mutations and which may pose a public health risk.

### VP1 informativeness

Polioviruses are among the fastest evolving RNA viruses and we observed that the rapid generation of genetic diversity in vaccinated children and their household and community contacts contains epidemiological information. Early evolution of OPV is largely clocklike, with variation in mutation rate across serotypes. The lack of a signal of clocklike evolution of OPV-2 and OPV-3 in vaccinated children likely reflects the shorter sampling period for vaccinated children; unvaccinated household and community contacts were infected later in the study by vaccine-derived viruses which had already passed through at least one host.

Our estimate of the OPV-1 VP1 substitution rate of 1.2 × 10^−4^ substitutions/site/day (including all samples) was similar to previous estimates of OPV-1 evolution 0.83 × 10^−4^ substitutions/site/day^20^ (estimated from three years of surveillance for acute flaccid paralysis) and faster than the estimate of the molecular clock for wild poliovirus type 1 evolution of 0.28 × 10^−4^ substitutions/site/day, estimated from a set of 31 viruses sampled over ten years^10^. The differences in substitution rate may reflect the different sampling durations, as shorter sampling durations often yield higher pathogen substitution rates estimates, likely because purifying selection has not yet removed transient deleterious mutations are still present prior to the effect of purifying selection^21,22^. It is also likely that Sabin vaccine viruses evolve more rapidly than wildtype viruses.

We observe moderate genetic structure in VP1 sequences, with increasing genetic divergence between samples from different communities compared to that sampled within the same community, household, or longitudinally within an individual. Yet VP1 divergence is not sufficient to make predictions about transmission linkage. Whole genome sequences and additional epidemiological information about timing of infection and contacts could improve inference of transmission networks.

### Reversion to virulence

The rapid reversion to virulence of Sabin vaccines is well documented^23–25^. While each serotype has key attenuating mutations in the IRES, additional mutations along the genome, including in the VP1 gene, stabilize the attenuated phenotype^26^. In OPV-2, a predictable evolutionary route towards virulence includes gatekeeper mutations including an IRES mutation, followed by a VP1 gene mutation, and a reversion in the 5’ UTR, followed by a wave of recombination events^27,28^.

We found a high frequency of attenuating substitutions in VP1, with substitutions appearing early after vaccination. The higher frequency of OPV-2 and OPV-3 reversion mutations we observed could reflect the greater observed genetic stability of OPV-1, which differs from the virulent wild-type progenitor virus by 59 substitutions, compared to OPV-2 and OPV-3 which are more closely related to the virulent ancestor^29^. Our group previously found that 97% of OPV-3 recipients shed viruses that had a site 472 reversion mutation within two weeks following vaccination^30,31^. The instability of OPV-3 has epidemiological consequences: OPV-3 causes the majority of vaccine-associated paralytic poliomyelitis (VAPP), paralysis attributable to the vaccine itself, followed by OPV-2 and then OPV-1^3^.

However, previous studies have reported heterogeneity at the OPV-3 C2493U attenuating mutation in vaccine stocks^32^. We did not have access to vaccine stocks and therefore cannot determine if the near-fixation of an attenuating mutation we observe was due to heterogeneity in the vaccine or *de novo* evolution in vaccinated children and subsequent onwards transmission.

### Identifying viruses of public health concern

Although the majority of OPV samples in our study collected were defined as Sabin-like viruses, rather than VDPVs, based on sequence divergence, many of the Sabin-like viruses had rapidly lost attenuating mutations at previously described positions. While VP1 sequence divergence may be useful in estimating duration of circulation or predicting transmission cluster, OPV circulating for short periods, that does not yet meet GPEI definitions for a cVDPV, can be virulent and can pose a public health risk in populations without prior immunity. For example, Sabin-like OPV-2 viruses, which were 3 SNPs distant to the Sabin virus VP1 sequence, were previously found to have caused a cluster of acute flaccid paralysis cases in an orphanage in Altai Region of Russia^33^. All outbreak genomes shared 15 substitutions compared to the Sabin virus, including the loss of attenuation mutations A481G and U2909C…

The global increase of cVDPV2 outbreaks has increased dramatically since 2016, when Sabin 2 was removed from the trivalent poliovirus vaccine^5^. Our results are consistent with previous work on the genetic instability of OPV and rapid reversion to virulence. Our finding that current definitions of cVDPVs may exclude virulent Sabin-like viruses underscores the urgency in heightened surveillance during and following vaccination campaigns.

## Data Availability

https://www.ncbi.nlm.nih.gov/genbank/

## Funding Information

This project was funded by the National Institutes of Health 5R21AI148810 to YM. KSW was funded by a Thrasher Early Career Award and a Stanford Child Health Research Institute postdoctoral fellowship.

## Supplementary Figures

**Figure S1.**
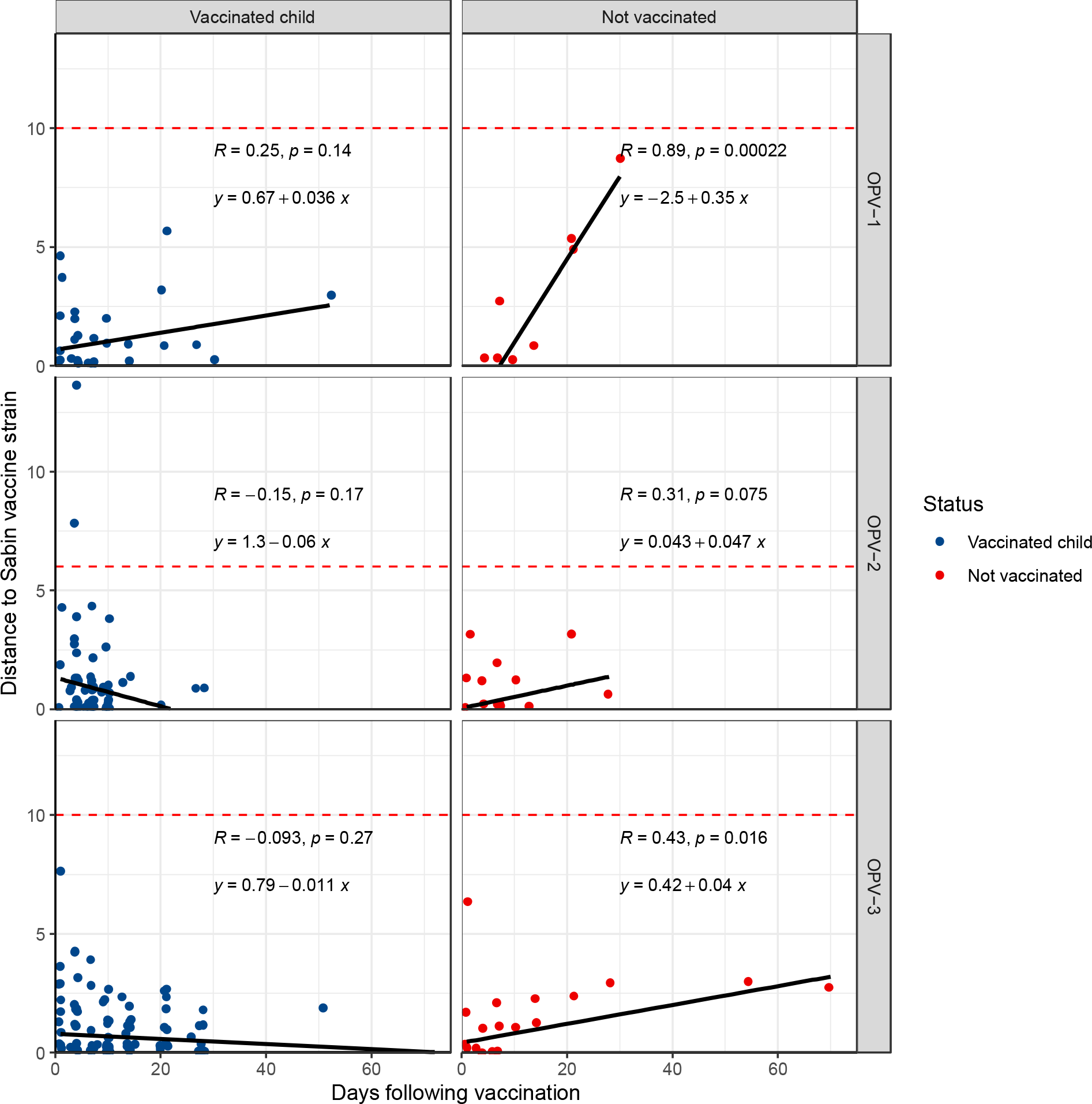
Measurable evolution in the OPV VP1 gene following vaccination campaigns when excluding attenuating mutations. Genetic distance of the OPV VP1 gene to the Sabin vaccine strain versus time from vaccination for vaccinated children (blue) and not vaccinated study participants (red), for each OPV serotype (a-c). Black lines and grey shading of a linear model for distance to the Sabin strain. Days following vaccination is measured as days from first vaccination (for vaccinated children), day from first household vaccination (for non-vaccinated household members), and days from first community vaccination (for non-vaccinated community members). Red dashed lines indicate the genetic distance threshold for a cVDPV: 10 SNPs distant from the Sabin vaccine for serotypes 1 and 3 and 6 SNPs distant for serotype 2.

## Notes

### Competing Interest Statement

The authors have declared no competing interest.

### Author Declarations

The study was approved by the Stanford University Institutional Review Board (Protocol #31546), the Comite de Etica, Bioseguridad e Investigacion of the Instituto Nacional de Salud Publica (CI: 1260, No. 1581), and the Instituto Veracruzano para la Formacion e Investigacion en Salud (SESVER/IVEFIS//SIS/DIB/0109/02014, classification 15S).

